# Assisting focused attention meditation through EEG-based alpha-theta cross-frequency neurofeedback

**DOI:** 10.1101/2024.09.05.24311404

**Authors:** Hendrik-Jan De Vuyst, Angeliki-Ilektra Karaiskou, Javier R. Soriano, Jelle Pergens, Ruth Op de Beeck, Katleen Van der Gucht, Filip Raes, Maarten De Vos, Carolina Varon, Kaat Alaerts

## Abstract

**Objectives:** Neurofeedback training involves real-time monitoring and self-regulation of neural activity. Neurofeedback training paradigms have been widely employed in the context of meditation. Interestingly, prior research revealed focused attention meditation to be associated with desynchronized, non-harmonic, cross-frequency relationships between alpha and theta rhythms, suggesting cross-frequency decoupling. However, the potential of training these brainwave patterns to assist meditative practices remains unexplored.

**Methods:** We assessed the trainability of non-harmonic alpha-theta cross-frequency relationships during focused attention meditation through EEG-neurofeedback training. Thirty individuals underwent 25 minutes of both experimental and sham training. During experimental training, participants received auditory feedback upon detection of non-harmonic alpha-theta brainwaves, whereas during sham training, feedback was unrelated to the measured brainwaves. Neural changes were assessed locally at training site Pz and globally across all scalp electrodes.

**Results:** Mixed model analyses showed a global, but not local, interaction effect between trainings over time, indicating that the incidence of non-harmonic alpha-theta relationships across the scalp increased during experimental training compared to sham training (*p* < 0.001). This effect persisted in the post-training resting-state recording (*p* = 0.004). Notably, these training-induced increases were associated with improvements in depressive mood state (*p* < 0.001). Furthermore, participants with a higher depressive mood state at baseline showed stronger training effects (*p* < 0.001).

**Conclusions:** Neurofeedback training can be used to upregulate non-harmonic alpha-theta cross-frequency relationships during focused attention meditation with durable post-training effects, particularly for those experiencing depressive mood symptoms. These findings lay the groundwork for investigating the effectiveness of multiple-session neurofeedback-assisted mindfulness training.

## Introduction

Mindfulness practices have gained widespread recognition as tools to support mental and physical health (Baminiwatta & Solangaarachchi, 2021; Kee et al., 2019; Zhang et al., 2021). While these practices vary in form, they are fundamentally rooted in the act of using one’s attention to observe the present moment with an attitude of non-judgmental acceptance and equanimity (Creswell et al., 2019; Kabat-Zinn, 2018). As such, the practice of mindfulness entails an enhanced awareness of one’s moment-to-moment experience, characterized by a temporary reduction in mind wandering (i.e., the tendency of the mind to drift off into spontaneous self-generated thought; Brandmeyer & Delorme, 2018; Feruglio et al., 2021; Turkelson et al., 2022). Given that mind wandering frequently involves negative self-referential content, its reduction is thought to be a key process through which mindfulness interventions positively impact mental health (Bortolla et al., 2022; Maltais et al., 2019; Vago & Zeidan, 2016).

The neural underpinnings of mindfulness meditation and its association with mind wandering have been extensively studied. In this regard, studies using electro-encephalography (EEG) have consistently underscored the relevance of alpha (8 – 14 Hz) and theta (4 – 8 Hz) band activity in these processes. Notably, alpha and theta band activity are intricately connected to working memory processes (Riddle et al., 2020). In the context of mindfulness meditation, meta-analyses point towards a simultaneous increase in both alpha and theta power (Lee et al., 2018; Lomas et al., 2015). At the same time, mind wandering, which is considered to be a departure from the meditative state, has been associated with a reduction in alpha power combined with an increase in theta power (Brabosczc & Delorme, 2011; Brandmeyer & Delorme, 2018; Dias da Silva et al., 2022; Kam et al., 2022; van Son et al., 2019a; van Son et al., 2019b). However, contrasting findings have been reported, suggesting that the relationship between mind wandering and changes in alpha and theta power may be more complex and context-dependent (Rodriguez-Larios et al., 2021).

Besides changes in the power of alpha and theta rhythms during mindfulness meditation and mind wandering, various studies have explored other EEG spectral parameters, including assessments of changes in the peak frequency (i.e., the frequency associated with the highest amplitude) of these rhythms. In this regard, studies have associated mindfulness practices with a decrease in alpha peak frequency (i.e., deceleration of alpha rhythm) and an increase in theta peak frequency (i.e., acceleration of theta rhythm; Cahn & Polich, 2006; Irrmischer et al., 2018; Kim et al., 2013; Saggar et al., 2012; Tripathi et al., 2022). Importantly, such transient shifts in peak frequencies may lead to different cross-frequency interactions by facilitating harmonic and non-harmonic relationships (Klimesch, 2018; Palva & Palva, 2018; Pletzer et al., 2010). This notion is rooted in the mathematical concept that neural oscillations can only achieve so-called full synchronization when their peak frequencies constitute harmonic 2:1 relationships (*f*_2_ = *f*_1_/2; e.g., *f*_1_ = 10 Hz and *f*_2_ = 5 Hz), thereby allowing a pattern of frequent and regular excitatory phase meetings (Klimesch, 2018). In contrast, non-harmonic relationships, involving irrational cross-frequency ratios, provide desynchronized states and preclude spurious, noisy background coupling (Klimesch, 2018; Palva & Palva, 2018). Notably, cross-frequency ratios based on the irrational phi ratio (∼1.6:1) have been associated with the highest physiologically possible level of desynchronization (*f*_2_ = *f*_1_/∼1.6; e.g., *f*_1_ = 10 Hz and *f*_2_ = 6 Hz; Klimesch, 2018; Palva & Palva, 2018; Pletzer et al., 2010).

The association between mindfulness meditation and changes in alpha-theta cross-frequency arrangements was first explored by Rodriguez-Larios et al. (2020) in a sample of experienced meditators. The authors observed a widespread linear decrease in the incidence of harmonic alpha-theta relationships when instructing participants to transition from cognitive task engagement to a resting state and eventually to breath-focus meditation (Rodriguez-Larios et al., 2020). This reduction in alpha-theta harmonization may suggest that mindfulness practices are linked with a neural environment characterized by a decreased demand for working memory processing. Indeed, given the crucial role of these rhythms in working memory, their synchronized activity may become less essential when the brain shifts into more restful states (Riddle et al., 2020). In a later study involving an experience sampling paradigm, Rodriguez-Larios and Alaerts (2021) observed a similar shift away from harmonic alpha-theta arrangements when instructing novice meditators to engage in breath-focus meditation. Importantly, these non-harmonic relationships would revert back into harmonic arrangements during episodes of mind wandering (Rodriguez-Larios & Alaerts, 2021). In light of these findings, the authors speculate that the harmonization of alpha and theta rhythms likely plays a key role in the process of mind wandering, which heavily relies upon working memory engagement (e.g., thinking about the past or future; anticipating or rescripting scenarios; Brandmeyer & Delorme, 2021; Rodriguez-Larios et al., 2021). Non-harmonic alpha-theta arrangements, on the other hand, may reflect a neural marker for the absence of such mind-wandering episodes and putatively signal a state of meditative awareness (Rodriguez-Larios & Alaerts, 2021; Senoussi et al., 2022; Wianda & Ross, 2019). This notion closely aligns with the suggestion that diminished working memory activity during mindfulness meditation may mirror its fundamental characteristic: a state of non-elaborative attentiveness to the present moment (Vago & Zeidan, 2016). Furthermore, recent studies show that oscillatory systems with a high incidence of non-harmonic cross-frequency arrangements are reflected by a higher ‘rest & relax’ parasympathetic drive, as assessed using heart rate variability, an established index of parasympathetic nervous system activity (Alaerts et al., 2021; Alaerts et al., 2024).

The notion that non-harmonic alpha-theta arrangements, suggestive of a neural idling state, closely correlate with the occurrence of meditative awareness and the absence of mind wandering presents new possibilities for both practitioners and researchers. Specifically, by drawing upon this association, exploring the potential of upregulating these specific brain rhythm configurations to assist meditative practices becomes feasible. In this regard, neurofeedback training emerges as a particularly promising method. Neurofeedback is a form of biofeedback that involves the real-time monitoring of brain activity, usually through EEG, and presenting these measurements in an understandable form back to the involved individual (Marzbani et al., 2016). This technique would allow individuals to observe their brain activity patterns as they engage in mindfulness meditation and learn to upregulate the occurrence of non-harmonic alpha-theta configurations to assist their meditative practice.

Evidence is emerging that neurofeedback training can indeed be used as a neural modality to assist meditative practices. For example, Chow et al. (2017) demonstrated how a 15-minute neurofeedback-assisted meditation training resulted in higher alpha power when compared to meditation alone. In a study by Brandmeyer and Delorme (2020), it was demonstrated that eight sessions of neurofeedback-assisted meditation training led to significant improvements in theta power and cognitive performance when compared to a control group receiving sham neurofeedback. Importantly, these studies focus on modulating individual brain rhythms and overlook the dynamic interactions of such rhythms across frequencies. At the same time, alpha-theta cross-frequency interactions appear to play key roles in the process of meditative awareness, or conversely, mind wandering. Consequently, the potential of cross-frequency neurofeedback training to modulate the interplay between alpha and theta brain rhythms to assist meditation practices remains a promising yet unexplored area.

A first step towards examining the potential of utilizing neurofeedback training to assist meditative practices is to assess the trainability of the underlying neural parameter; that is, a non-harmonic alpha-theta configuration characterized by the irrational ∼1.6:1 cross-frequency ratio (Pletzer et al., 2010). To this end, we have designed the current within-subject neurofeedback study involving 30 participants taking part in two training protocols: (1) an experimental, auditory 25-minute neurofeedback training targeting the upregulation of non-harmonic alpha-theta cross-frequency interactions during focused attention meditation; and (2) a sham training where auditory feedback signals are unrelated to the recorded brain activity. We hypothesized that, compared to sham training, participants following the experimental training would show a stronger increase in the occurrence of non-harmonic alpha-theta ratios approximating ∼1.6:1 across the training trials. Further, to examine whether the training effects were retained outside the explicit training context, a resting-state EEG recording was obtained before and after each training run. Here, we hypothesized that, compared to sham training, participants following the experimental training would show a stronger increase in the occurrence of non-harmonic alpha-theta ratios approximating ∼1.6:1 as measured from the pre-training resting-state recording to the post-training resting-state recording. Finally, we explored possible brain-behavior associations by examining whether upregulating alpha-theta non-harmonicity was related to changes in self-reported psychological and physiological states.

## Methods

### Participants

This study included a total of 30 participants. Power calculations indicated that a sample size of 27 participants would allow detecting a medium effect of training (*d* = 0.50) with α set at 0.05 and power set at 0.80. Three additional participants (i.e., +10%) were included to anticipate missing or erroneous data. Inclusion criteria comprised being aged between 18 and 35 years old, not taking any psychopharmacological medication (e.g., antidepressants, anxiolytics, antipsychotics), and having no prior experience with meditation practices. The final sample consisted of 8 men and 22 women aged between 18 and 34 years old (*M* = 22.37; *SD* = 2.86). Participants were recruited through flyers distributed at university campuses and on social media. All participants provided written informed consent prior to participation. At the end of the study, participants were remunerated for their participation at a rate of €10 per hour. The study was preregistered on the OSF platform (https://osf.io/z9wck/?view_only=57e15a9cd31f489b8a366cd925960195). The study protocol was approved by the Social and Societal Ethics Committee (SMEC) of KU Leuven (G-2022-5975).

### Procedure

The study involved a within-subjects design in which participants took part in an experimental neurofeedback training and a sham training, presented in a randomized sequence. In other words, half of the participants began with the experimental training, while the other half started with the sham training. Prior to the training, participants completed self-report assessments on emotional distress, mindfulness skills, and mood state. At the end of the training conditions, participants again self-reported their mood state. During the training, participants sat in a dimly lit room in front of a computer screen and were fitted with an EEG cap to measure their brain activity (as well as other physiological sensors; not included in the current report). Next, participants received a brief introduction to the rationale and structure of the study and then received earphones to individually adjust the volume level of the auditory stimuli played throughout the training.

Each training condition lasted approximately 25 minutes and started and ended with a 2-minute resting-state recording. During these resting-state recordings, participants were instructed to sit still with closed eyes and let their thoughts wander freely without intentionally thinking of anything specific. During the neurofeedback training, participants were presented with ten 2-minute neurofeedback trials. During these trials, participants were again instructed to close their eyes and refrain from moving. Additionally, participants received instructions to engage in a meditative exercise involving focused attention towards an anchor point. This type of meditative instruction required participants to intentionally direct their attention on a particular object and gently refocus attention whenever distractions arose (Lutz et al., 2008). In this context, the focal point of attention was directed towards the sensation of sitting, specifically concentrating on the area where the buttocks come into contact with the seating surface. During the experimental training, continuous EEG recordings were performed to calculate the auditory feedback signal (i.e., a woodblock percussive tapping sound). During sham training, feedback sounds were not contingent upon the participant’s measured brain activity. Considering current models of neurofeedback learning through operant conditioning, participants were instructed to refrain from consciously attempting to adopt strategies to reproduce as many feedback sounds as possible but to merely perceive these sounds as signals that their brain activity was aligning with the targeted neural state (Sitaram et al., 2017). Indeed, it has been shown that deliberate efforts to manipulate neurofeedback can inhibit the process of meditation, which fundamentally revolves around non-goal-oriented behavior (Prestel et al., 2019). Following every two training trials, participants were asked to indicate their subjective experience of agitation, sleepiness, relaxation, and focus during those trials, on a Likert scale ranging from 1 (*not at all*) to 5 (*completely*) using a numerical keypad.

Next to the feedback sounds, all recording intervals were accompanied by a constant background sound (i.e., a sustained echo of a bell) to mask any external distracting noises, as well as a start-and-stop sound to indicate when participants should open or close their eyes. At all times, instructions were also displayed on the computer screen to guide participants through the training.

## Measures

### Self-Report Questionnaires

Mindfulness skills were assessed using the Mindful Attention Awareness Scale (MAAS; Brown & Ryan, 2003). Levels of emotional distress were measured using the Depression, Anxiety, and Stress Scale (DASS-21; Lovibond & Lovibond, 1995). Mood state was measured using 11 selected items (focusing on tiredness, vigilance, agitation, and depression) of the Profile Of Mood States questionnaire (POMS; Grove & Prapavessis, 1992).

### EEG Recordings

The Nexus-32 system and BioTrace software (V2018A1; MindMedia, the Netherlands) were used to obtain EEG recordings. The EEG data was recorded using a 22-electrode cap (including one ground electrode and two reference electrodes on the mastoids) positioned according to the 10-20 system (Mindmedia, the Netherlands). Additionally, four pre-gelled electro-oculographic electrodes were placed around the eyes to detect vertical and horizontal eye movements (Kendall, Germany). Electrode paste (Nuprep) was used to reduce electrode impedances. The measured EEG signal was amplified using a unipolar amplifier with a sampling rate of 1024 Hz. OpenVibe software was used for data quality checks during sensor placement and for data monitoring during the experiment. The recorded data was streamed to Matlab (2019a) and recorded through Lab Stream Layer (Renard et al., 2010).

### Online Pre-Processing

Throughout the neurofeedback training trials, continuous EEG recordings were captured at the midline electrode Pz, chosen for its favorable potential in modulating alpha and theta rhythms (e.g., Boynton , 2001). The recorded data were preprocessed in real-time at non-overlapping 1-second intervals using the EEGLAB toolbox in Matlab (Delorme & Makeig, 2004). First, a high-pass filter with a 1 Hz cut-off frequency was applied to the raw EEG data to attenuate low-frequency noise using the *pop-eegfiltnew* function. Next, a 50 Hz notch filter was used to suppress the line noise originating from the power supply (5^th^ order Butterworth filter with cut-off frequencies of 49 – 51 Hz). Finally, the EEG data was downsampled from 1024 Hz to 256 Hz using the *pop_resample* function to reduce further data processing time.

### Offline Pre-Processing

The raw EEG data was preprocessed in EEGLAB. First, using the *pop_eegfiltnew* function, a high-pass filter with a 1 Hz cut-off frequency was applied to attenuate low-frequency noise. Next, a 50 Hz notch filter was used to suppress powerline interferences (5^th^ order Butterworth filter with cut-off frequencies of 49 – 51 Hz). Subsequently, a low-pass filter at 40 Hz was applied to attenuate high-frequency noise using the *pop_eegfiltnew* function. Following this, any channels with poor or absent signals were removed using the *clean_flatlines* and *clean_channels* functions of EEGLAB with a threshold of 0.5. Next, the removed channels were reconstructed using spherical interpolation using the *pop_interp* function. Then, the data was re-referenced using the *pop_reref* function to the standard average reference. Subsequently, an independent component analysis was performed using the *pop_runica* and the *iclabel* algorithm to reject noisy components associated with muscle, eye, heart, or channel noise. Finally, all data was downsampled from 1024 Hz to 256 Hz using the *pop_resample* function.

### Peak Detection and Cross-Frequency Ratio Calculation

The Short-term Fast Fourier Transform, as implemented in the function *spectrogram* of MATLAB, was applied to each 1-second batch of EEG data, employing a sliding Hanning window of 1-second length with a 91% overlap between consecutive windows. Consequently, a power spectral density plot with a frequency resolution of 0.1 Hz was generated. Next, transient peak frequencies within the theta (4 – 8 Hz) and alpha (8 – 14 Hz) frequency bands were identified using the *findpeaks* function (Figure 1A). Using this algorithm, data samples larger than their two neighboring samples were identified as local peaks within the specified frequency ranges. If multiple peaks were identified within one frequency band, the peak with the highest amplitude was used for further analysis. Based on the identified peak frequencies, the numerical ratio of the alpha and theta peaks (peak-frequency_alpha_ / peak-frequency_theta_) was calculated for each epoch and rounded to the first two decimal places (e.g., 8.9 Hz / 5.5 Hz = 1.62; figure 1B). During experimental training, participants received discrete auditory feedback in the form of a woodblock percussive tapping sound. This feedback was given for each 1-second epoch where the cross-frequency ratio approximated the non-harmonic ∼1.6:1 ratio; that is, within the range of 1.5:1 to 1.7:1, to ensure an adequate amount of feedback signals (∼20%), as guided by pilot studies (see Figure 1C). During sham training, feedback sounds were not contingent on the participant’s measured brain activity but were set to approximate the frequency expected during the experimental training sessions.

**Figure 1.**
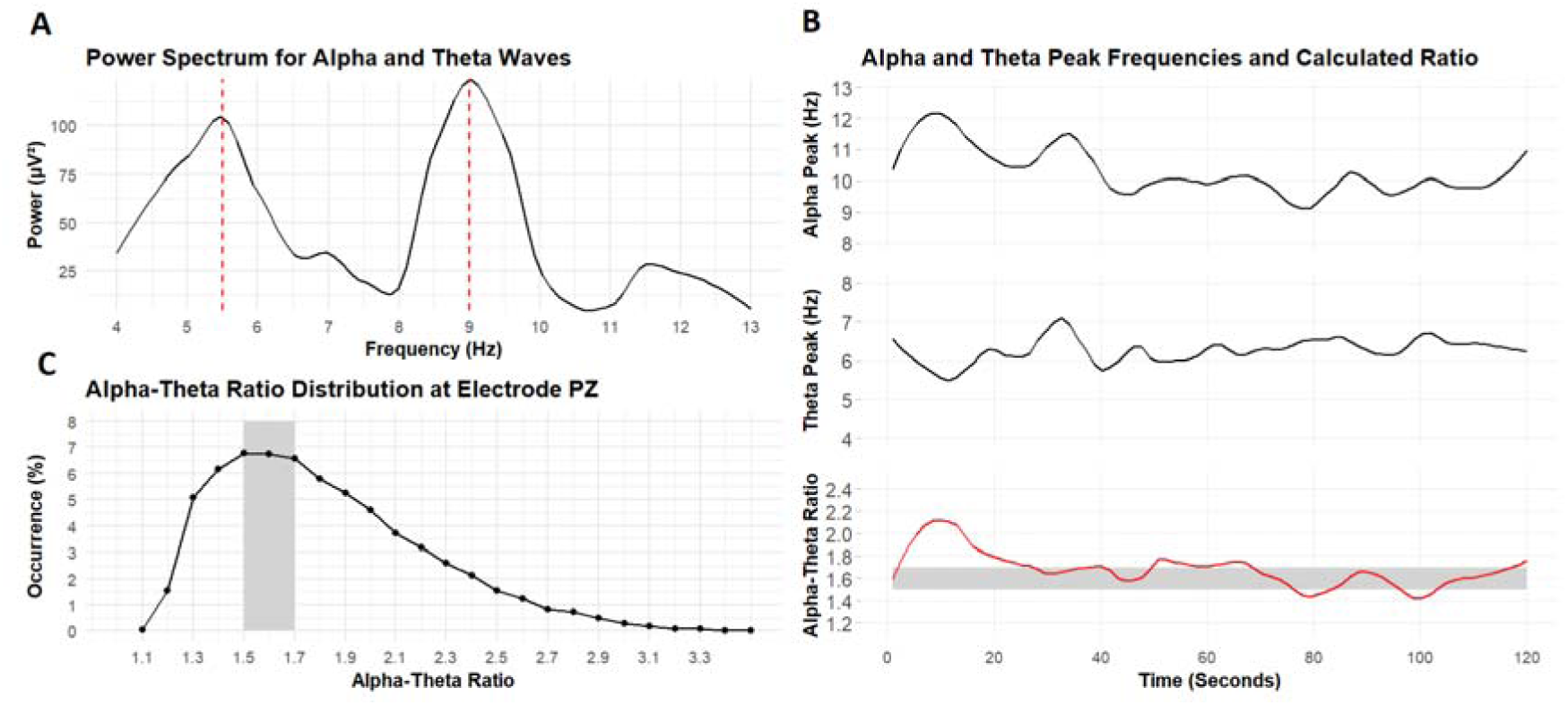
Calculation of alpha-theta cross-frequency ratios. Note. Panel A illustrates the frequency spectrum of an exemplary epoch where the identified alpha and theta peak frequencies exhibit a non-harmonic (∼1.6:1) cross-frequency relationship. Panel B depicts the instantaneous frequencies of alpha and theta rhythms, along with their numerical ratio over a 120-second timeframe (i.e., one training trial), for an exemplary subject at electrode Pz. Grey areas highlight epochs where alpha-theta ratios fall within the non-harmonic range centered around 1.6:1 (i.e., 1.5:1 to 1.7:1). Panel C displays the overall distribution of alpha-theta cross-frequency ratios recorded at electrode Pz across all subjects (recorded during the experimental training run), with grey areas indicating the proportion of ratios within the non-harmonic range centered around 1.6:1 (i.e., from 1.5:1 to 1.7:1).

## Statistical analyses

### Local Training Effects

To examine changes in the incidence of the non-harmonic alpha-theta ratio range throughout the training trials, a linear mixed effects modeling approach was adopted, with the incidence of the non-harmonic alpha-theta ratio range at training electrode Pz inserted as the dependent variable. The factors ‘Trial’ (trials 1 to 10), ‘Training’ (experimental vs. sham), and ‘Run’ (first run vs. second run) were included as fixed factors, whereas ‘Subject’ was included as a random factor.

Examinations of changes in the incidence of the non-harmonic alpha-theta ratio range at training electrode Pz from the pre-training to the post-training resting-state recordings were similarly performed using a linear mixed effects modeling approach with the factors ‘Time Point’ (pre-training vs. post-training), ‘Training’ (experimental vs. sham), and ‘Run’ (first run vs. second run), included as fixed factors and ‘Subject’ as a random factor.

### Global Training Effects

Following these targeted analyses, a global model was introduced by including ‘Electrode’ as a fixed factor to explore the training’s impact on the incidence of the non-harmonic alpha-theta ratio range across all 19 scalp electrodes (not pre-registered). This approach aligns with findings from previous studies, which reported that changes in the incidence of non-harmonic alpha-theta ratios were not localized to specific brain regions but rather broadly distributed across the scalp (Rodriguez-Larios et al., 2020; Rodriguez-Larios et al., 2020; Rodriguez-Larios & Alaerts, 2021).

### Brain-Behavior Associations

To explore associations between global changes in the non-harmonic alpha-theta ratio range and behavioral assessments, Spearman correlation analyses were conducted. First, we examined the relationship between initial baseline assessments of emotional distress, mindfulness skills, and mood state and the concurrently assessed incidence of the non-harmonic alpha-theta ratio range. Subsequently, we investigated how these baseline assessments predicted neurofeedback trainability, which was defined as the change in the incidence of the non-harmonic alpha-theta ratio range from the pre-experimental to post-experimental resting-state recordings. Next, we assessed the association between changes in the non-harmonic alpha-theta ratio range (from the first to the last resting-state recording) and changes in simultaneously assessed mood states. Lastly, we explored the relationship between changes in self-reported psychological and physiological states (i.e., levels of sleepiness, relaxation, focus, and agitation) during training and corresponding changes in the incidence of the non-harmonic alpha-theta ratio range. These outcomes were operationalized as the average reported levels during the first two training trials compared to the average reported levels during the last two training trials of each training.

### Spatial and Ratio Specificity

The spatial and ratio specificity of the changes in the incidence of the non-harmonic alpha-theta ratio range following the experimental training was tested using a cluster-based permutation testing approach. Specifically, changes in ratio occurrences between the first and last training trial were assessed by performing paired samples t-tests for all (electrode, ratio)-pairs. The pairs exceeding a threshold (*q* = .05) were then clustered on the basis of spatial and ratio adjacency, and cluster-level statistics were calculated as the sum of the *t*-values within every cluster. A permutation method was used to estimate the significance probability at the cluster level by constructing the null distribution of the cluster values through 10,000 random permutations. Finally, the observed values were tested against the (1 − α)th percentile of the null distribution. All statistical analyses were conducted utilizing SPSS (Version 26.0, IBM Corp.) and Matlab (Version 2023a, The Mathworks Inc.), with the significance threshold set at an alpha level of .05.

## Results

### Trainability of Non-Harmonic Alpha-Theta Cross-Frequency Interactions Across Training Trials

#### Neurofeedback-Induced Changes at Training Electrode Pz

Linear mixed-effects analyses revealed no significant main or interaction effects for the factor Training (see Table 1 for an overview of the main test statistics). This finding indicates that, across training trials, there were no overall differences between the experimental and sham training conditions in the incidence of the non-harmonic alpha-theta ratio range at the Pz training site (Figure 2A). Also, no main or interaction effects were revealed for the factors Trial or Run, indicating no differential changes across trials or training runs (Figure 2A).

**Figure 2.**
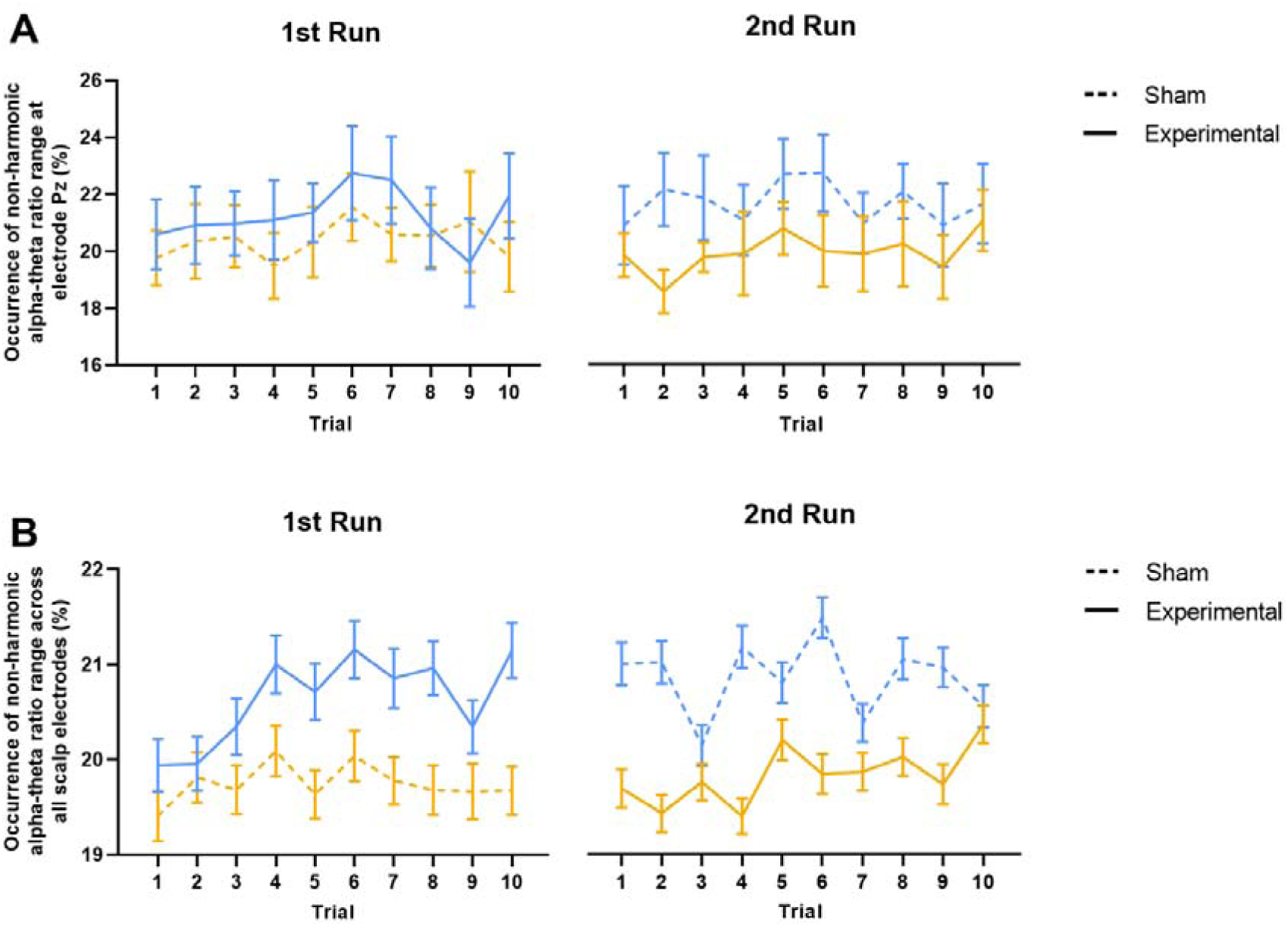
Changes in the incidence of the trained non-harmonic alpha-theta ratio range throughout the training. Note. Training-induced changes in the occurrence of the non-harmonic alpha-theta ratio range are visualized across trials, separately for each training condition (sham, experimental) and chronologically by training run (1 run, 2 run). Participants receiving the experimental training during the first run (and sham training during the second run) are visualized in blue (n = 15). Participants receiving the sham training during the first run (and experimental training during the second run) are visualized in orange (n = 15). Panel A visualizes changes observed at the training electrode Pz, while Panel B visualizes changes across all scalp electrodes. Vertical error bars represent the standard error of the mean.

**Table 1.** Main results of the linear mixed-effects model: Incidence of the trained non-harmonic alpha-theta ratio range across training trials.

| Predictor | Sum of<br>Squares | Mean<br>Squares | $DF_{Num}$ | $DF_{Den}$ | $F$ | $p$ |
| --- | --- | --- | --- | --- | --- | --- |
| **At Electrode Pz** |  |  |  |  |  |  |
| Trial | 0.013 | 0.001 | 9 | 532 | 0.879 | 0.544 |
| Training | 0.003 | 0.003 | 1 | 532 | 1.749 | 0.187 |
| Run | 0.000 | 0.000 | 1 | 532 | 0.071 | 0.789 |
| Trial x Training | 0.008 | 0.001 | 9 | 532 | 0.545 | 0.842 |
| Training x Run | 0.002 | 0.002 | 1 | 28 | 1.510 | 0.230 |
| Trial x Training x Run | 0.004 | 0.000 | 9 | 532 | 0.306 | 0.973 |
| **Across All Scalp Electrodes** |  |  |  |  |  |  |
| Trial | 0.060 | 0.007 | 9 | 10612 | 4.428 | < 0.001 |
| Training | 0.014 | 0.014 | 1 | 10612 | 9.388 | 0.002 |
| Channel | 0.331 | 0.018 | 18 | 10612 | 12.263 | < 0.001 |
| Run | 0.089 | 0.089 | 1 | 10612 | 59.408 | < 0.001 |
| Trial x Training | 0.063 | 0.007 | 9 | 10612 | 4.669 | < 0.001 |
| Training x Run | 0.002 | 0.002 | 1 | 28 | 1.347 | 0.256 |
| Trial x Training x Run | 0.029 | 0.003 | 9 | 10612 | 2.121 | 0.025 |

#### Neurofeedback-Induced Changes Across Electrodes

Using a global linear mixed-effects analysis including all 19 electrodes, we observed significant main effects of both Training, *F*(1, 10612) = 9.388, *p* = 0.002, and Trial, *F*(9, 10612) = 4.428, *p* < 0.001, as well as a significant Training × Trial interaction effect, *F*(9, 10612) = 4.669, *p* < 0.001 (see Table 1 for an overview of the main test statistics). This interaction effect signified that the trajectory of changes over trials in the incidence of the non-harmonic alpha-theta ratio range across the scalp was significantly different between the experimental and sham training. Post-hoc exploration of the interaction effect indicated that, during the experimental training, the incidence of alpha-theta non-harmonicity significantly increased from the first (*M* = 0.199, *SE* = 0.005) to the last (*M* = 0.210, *SE* = 0.005) trial, *t*(10612) = -4.555, *p* < 0.001, whereas during the sham training, the incidence remained unchanged from the first (*M* = 0.205, *SE* = 0.005) to the last (*M* = 0.204, *SE* = 0.005) trial, *t*(10612) = 0.682, *p* = 1.00.

Note, however, that also significant interaction effects with the factor Run were evident, including the three-way Training × Trial × Run interaction effect, *F*(9, 10612) = 2.121, *p* = 0.025, indicating that the aforementioned Training x Trial effect was different between the first and second runs. Specifically, during the first training run, participants receiving the experimental training showed a significant increase, *t*(10612) = -3.732, *p* = 0.0374, in the incidence of the non-harmonic alpha-theta ratio range from Trial 1 (*M* = 0.199, *SE* = 0.007) to Trial 10 (*M* = 0.211, *SE* = 0.007), whereas participants receiving the sham training showed no statistically significant changes, *t*(10612) = -0.832, *p* = 1.00, from Trial 1 (*M* = 0.194, *SE* = 0.007) to Trial 10 (*M* = 0.197, *SE* = 0.007). For the second training run, while no significant changes were detected in either group, participants who first received the experimental neurofeedback training showed a retention of the trained non-harmonic state from the first to the second run. More specifically, participants first receiving the experimental training retained an overall higher incidence of non-harmonicity during their entire subsequent sham training run (Figure 2B).

### Transferability of the Upregulated Non-Harmonic Cross-Frequency Interactions to Post-Training Resting-State Recording

#### Changes During Resting-State Recording at Training Electrode Pz

Linear mixed-effects analyses at electrode Pz revealed no significant main or interaction effects for the factor Training (see Table 2 for an overview of the main test statistics). This finding indicates that there were no overall differences between experimental and sham training conditions, nor were there any differential changes in the incidence of the non-harmonic alpha-theta ratio range from pre-training to post-training resting-state recordings in either group (Figure 3A).

**Figure 3.**
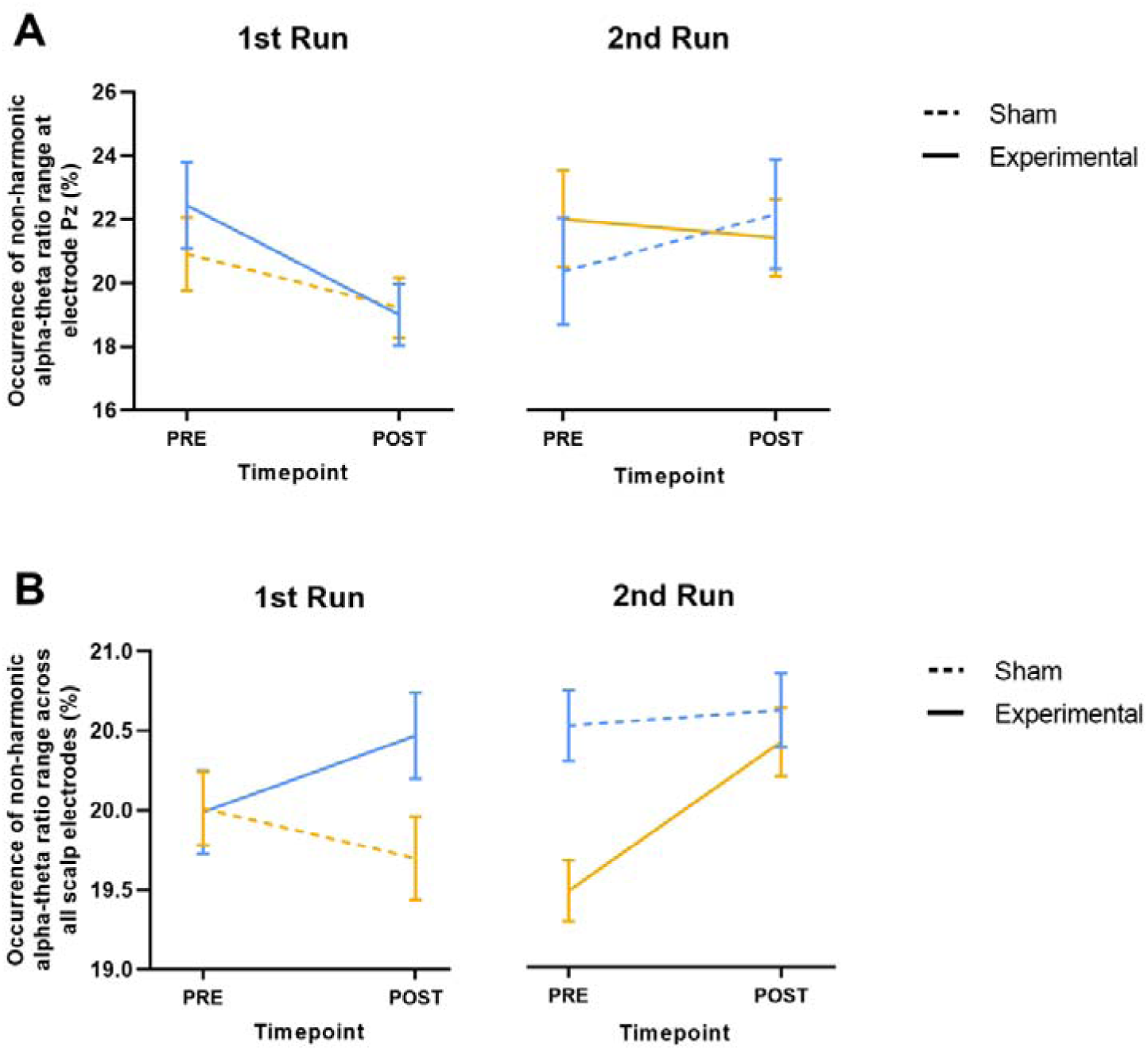
Resting-state changes in the incidence of the trained non-harmonic alpha-theta ratio range. Note. Changes in the occurrence of the non-harmonic alpha-theta ratio range are visualized from pre-intervention to post-intervention resting-state recordings, separately for each training condition (sham, experimental) and chronologically by training run (1 run, 2 run). Participants receiving the experimental training during the first run (and sham training during the second run) are visualized in blue (n = 15). Participants receiving the sham training during the first run (and experimental training during the second run) are visualized in orange (n = 15). Panel A visualizes changes observed at the training electrode Pz, while Panel B visualizes changes across all scalp electrodes. Vertical error bars represent the standard error of the mean.

**Table 2.** Main results of the linear mixed-effects model: Incidence of the trained non-harmonic alpha-theta ratio range from the pre-intervention resting-state recording to the post-intervention.

| Predictor | Sum of<br>Squares | Mean<br>Squares | $DF_{Num}$ | $DF_{Den}$ | $F$ | $p$ |
| --- | --- | --- | --- | --- | --- | --- |
| **At Electrode Pz** |  |  |  |  |  |  |
| Time point | 0.003 | 0.003 | 1 | 84 | 2.108 | 0.150 |
| Training | 0.001 | 0.001 | 1 | 84 | 0.501 | 0.481 |
| Run | 0.000 | 0.000 | 1 | 84 | 0.000 | 0.989 |
| Timepoint x Training | 0.002 | 0.002 | 1 | 84 | 1.611 | 0.208 |
| Training x Run | 0.000 | 0.000 | 1 | 28 | 0.018 | 0.895 |
| Timepoint x Training x Run | 0.000 | 0.000 | 1 | 84 | 0.002 | 0.962 |
| **Across All Scalp Electrodes** |  |  |  |  |  |  |
| Time point | 0.008 | 0.008 | 1 | 2100 | 5.214 | 0.023 |
| Training | 0.002 | 0.002 | 1 | 2100 | 1.585 | 0.208 |
| Channel | 0.052 | 0.003 | 18 | 2100 | 1.992 | 0.008 |
| Run | 0.016 | 0.016 | 1 | 2100 | 10.977 | < 0.001 |
| Time point x Training | 0.012 | 0.012 | 1 | 2100 | 8.389 | 0.004 |
| Training x Run | 0.001 | 0.001 | 1 | 28 | 0.415 | 0.525 |
| Time point x Training x Run | 0.000 | 0.000 | 1 | 2100 | 0.180 | 0.672 |

#### Changes During Resting-State Recording Across Electrodes

Linear mixed-effects analyses examining changes in alpha-theta non-harmonicity across the entire scalp between the resting-state recordings showed a significant main effect of Time point, *F*(1, 2100) = 5.214, *p* = 0.023, as well as a significant Training x Time point interaction effect, *F*(1, 2100) = 8.389, *p* = 0.004 (see Table 2 for an overview of the main test statistics). This result indicates differential changes in the occurrence of the trained non-harmonic alpha-theta ratio range from the pre-training to the post-training resting-state recordings between the experimental and sham training runs (Figure 3B). Post-hoc exploration of this interaction effect showed a significant increase in the incidence of the non-harmonic alpha-theta ratio range across the scalp from the pre-experimental (*M* = 0.198, *SE* = 0.005) to the post-experimental (*M* = 0.206, *SE* = 0.005) resting-state recording *t*(2100) = - 3.663, *p* < 0.001. In contrast, no changes were evident from the pre-sham (*M* = 0.205, *SE* = 0.005) to the post-sham (*M* = 0.204, *SE* = 0.005) resting-state recording *t*(2100) = 0.433, *p* = 0.665. Together, this observation of a sustained higher incidence of the non-harmonic alpha-theta ratios during the resting-state recording following the experimental training provides additional support for the notion that the training effects are retained and likely transferable outside the active training context. Finally, note that while a significant main effect of Run was evident, no significant two- or three-way interactions with the factors Training or Time point were revealed, indicating that the observed training-induced changes from the pre-training resting-state recording to the post-training resting-state recording were similar across both training runs.

### Associations Between Behavioral Assessments and Non-Harmonic Cross-Frequency Interactions

#### Assessment of Changes in Mood States During and Following Training

Assessments of changes in mood states (as assessed using the POMS) over the course of the entire recording session showed that, overall, participants reported a significant improvement in feelings of agitation, *t*(29) = 3.039, *p* = 0.005, vigilance, *t*(29) = 5.943, *p* < 0.001, and depressive mood states, *t*(29) = 2.983, *p* = 0.005, irrespective of whether they received the experimental or sham training first. Furthermore, correlation analyses showed that, particularly for the improvement in self-reported depressive mood (i.e., changes from the start to the end of the recording session), a negative association with changes in alpha-theta non-harmonicity was evident, ρ(28) = -.528, *p* < 0.001. This finding indicates that participants who exhibited stronger reductions in self-reports of depressive mood states showed greater increases in the incidence of the non-harmonic alpha-theta ratio range from the first to the last resting-state recording (Figure 4C; see supplementary Table 2 for all test statistics). During the training trials, no significant associations were found between changes in the incidence of the alpha-theta non-harmonic ratio range and changes in self-reported levels of sleepiness, relaxation, agitation, or focus (see supplementary Table 3 for all test statistics).

**Figure 4.**
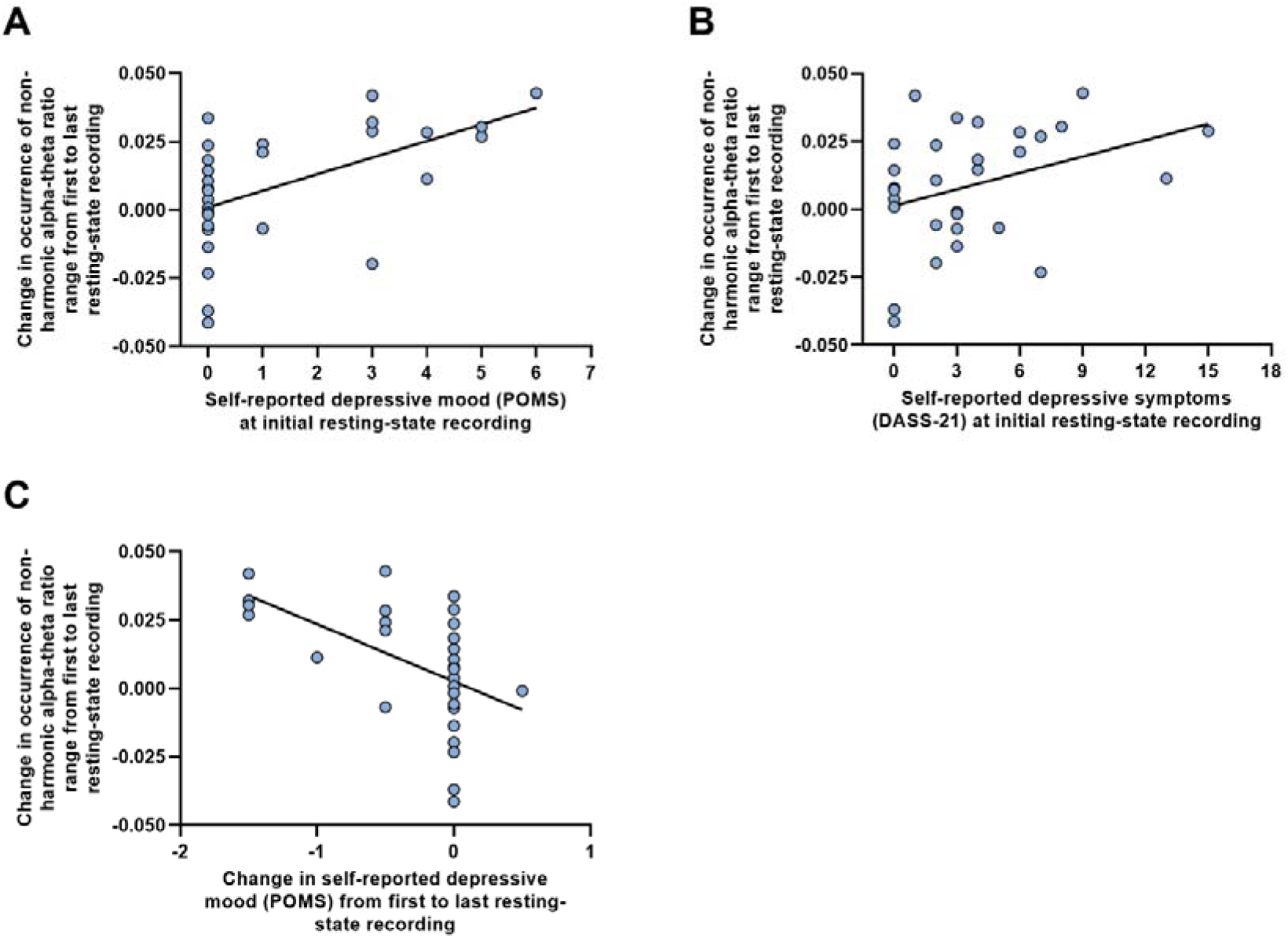
Brain-behavior associations between changes in alpha-theta non-harmonicity and self-reported behavior during rest and training. Note. Panel A visualizes the association between self-reported depressive mood state (depression subscale of the POMS) at the initial resting-state recording and changes in the incidence of the non-harmonic alpha-theta ratio range from the first resting-state recording to the last resting-state recording. Note that higher scores on the POMS represent a higher depressive mood state. Panel B visualizes the association between self-reported depressive symptomatology (depression subscale of the DASS-21) at the initial resting-state recording and changes in the incidence of the non-harmonic alpha-theta ratio range from the first resting-state recording to the last resting-state recording. Note that higher scores on the DASS-21 represent higher general depressive symptomatology. Panel C visualizes the association between changes in self-reported depressive mood state (depression subscale of the POMS) from the first resting-state recording to the last resting-state recording and changes in the simultaneously assessed incidence of the non-harmonic alpha-theta ratio range. Note that higher scores on the POMS represent a higher depressive mood state.

#### Assessment of Baseline Emotional Distress, Mindfulness Skills, and Mood States

Overall, baseline behavioral assessments of emotional distress, mindfulness skills, and mood states were not significantly associated with the concurrently assessed incidence of non-harmonic alpha-theta interactions. However, correlational analyses revealed that individuals with a higher baseline depressive mood state (Figure 4A), as assessed by the depression subscale of the POMS, and those with a higher baseline general depressive symptomatology (Figure 4B), as assessed by the depression subscale of the DASS-21, exhibited greater neurofeedback trainability (ρ(28) = .574, *p* < .001, and ρ(28) = .376, *p* = .040, respectively). Trainability was defined as the increase in the incidence of the non-harmonic alpha-theta ratio range from pre-training to post-training. See supplementary Table 1 for all test statistics.

### Spatial and Ratio Specificity of the Upregulated Alpha-Theta Ratios

We employed a cluster-based, non-parametric randomization approach to evaluate the effects of the experimental training on the prevalence of cross-frequency ratios between alpha and theta rhythms at all different electrodes (Figure 5). Following correction for multiple testing, results indicate a significant increase in the occurrence of a cluster of ratios, ranging between 1.3:1 and 1.6:1, at electrodes FP1, FP2, FZ, F4, and F8 when comparing Trial 10 to Trial 1. At the same time, a significant decrease was noted in the occurrence of ratios ranging between 2.1:1 and 2.2:1 at electrodes F3 and FZ. The findings highlight a shift in ratios, particularly at the frontal electrodes, moving from ratios approximating the harmonic 2.0:1 ratio to ratios encompassing the non-harmonic 1.6:1 ratio throughout the trials.

**Figure 5.**
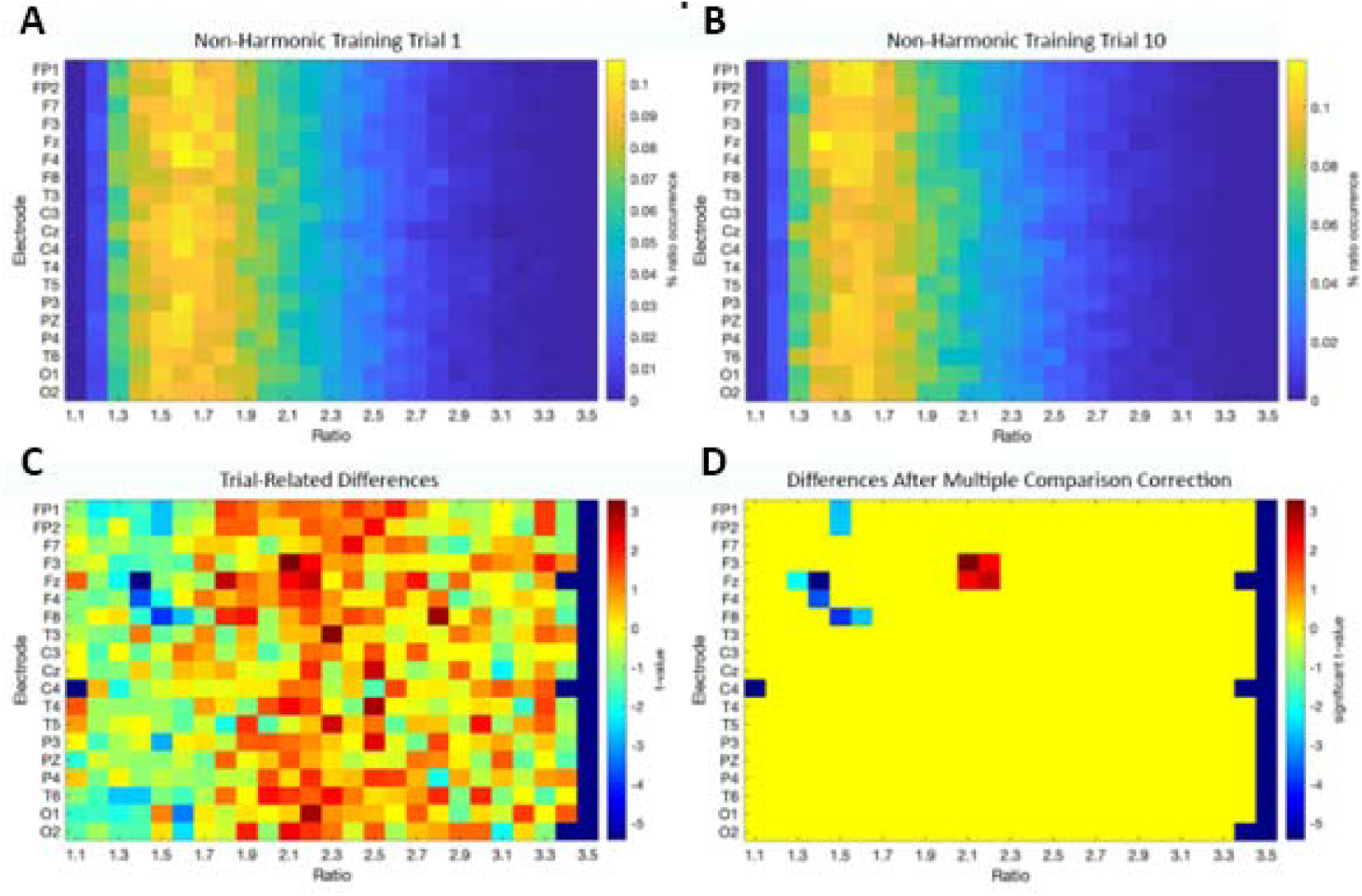
Differences in the incidence of distinct alpha-theta cross-frequency ratios between trials 1 and 10 of the experimental training. Note. Panel A visualizes the distribution of the percentage of each ratio occurrence for Trial 1 of the experimental training, separately for each electrode (y-axis) and cross-frequency ratio (x-axis; ratios ranging between 1.1 and 3.5). Panel B visualizes the distribution of the percentage of each ratio occurrence for Trial 10 of the experimental training, separately for each electrode (y-axis) and cross-frequency ratio (x-axis; ratios ranging between 1.1 and 3.5). Panel C visualizes the colormap of t-values derived from the cluster-based permutation analysis estimating the training-related effect (paired sample t-test; trial 10 vs. trial 1), separately for each electrode (y-axis) and cross-frequency ratio (x-axis). Permutation analyses were performed for ratios ranging between 1.1 and 3.5. Panel D shows the map of t-values masked for significance after the multiple comparison correction of the cluster-based permutation (p < 0.025).

## Discussion

The potential of using neurofeedback training to assist meditative practices has captured growing interest (Tarrant, 2020). At the same time, recent studies provided initial indications of an association between non-harmonic ratios among alpha and theta rhythms and states of focused attention meditation (Rodriguez-Larios et al., 2020; Rodriguez-Larios et al., 2020; Rodriguez-Larios & Alaerts, 2021). The current study builds on these premises as we sought to investigate the effectiveness of neurofeedback training to upregulate non-harmonic alpha-theta cross-frequency interactions during focused attention meditation, which may aid in assisting meditative practices.

### Trainability of Non-Harmonic Alpha-Theta Cross-Frequency Interactions

Results show that the incidence of the non-harmonic alpha-theta ratio range increased across the training trials during the experimental training but not during the sham training. This training effect was, however, only evident for the global, whole-scalp analysis, and thus not for the local analysis focusing only on the trained Pz electrode. This result points towards potential whole-brain trainability as it suggests that the experimental training had a more widespread impact on the incidence of the non-harmonic alpha-theta ratio range rather than being confined to the trained Pz electrode. This broader distribution of neurofeedback effects is consistent with the understanding that EEG signals at any given electrode likely represent a mixture of activity recorded from various neural source generators, i.e., due to volume conductive effects, thereby capturing the collective activity of underlying brain regions (White et al., 2014). It can therefore be expected that the effects of training at a single electrode will likely influence EEG dynamics in a more spatially distributed way (White et al., 2014). Furthermore, this finding is in accordance with previous studies assessing alpha-theta cross-frequency ratios in the context of mindfulness meditation, which reported that changes in the incidence of such ratios were not localized to specific brain regions but rather broadly distributed across the scalp (Rodriguez-Larios et al., 2020; Rodriguez-Larios et al., 2020; Rodriguez-Larios & Alaerts, 2021).

In the second training run, there were no significant changes in the incidence of the non-harmonic alpha-theta ratio range among participants, neither in the experimental training nor sham training. Here, the absence of a significant increase during the experimental training might be attributed to the effects of the preceding sham training. Specifically, the randomness and inconsistency in feedback timing during the sham training, coupled with misaligned communication about its significance (namely, the approximation of a meditative state), may have introduced elements of confusion. This confusion may then have interfered with the participants’ ability to effectively engage in their subsequent experimental training run, thereby undermining its effectiveness. In view of this, future neurofeedback training studies may be urged to consider parallel designs instead of cross-over designs to avoid potential interference or carry-over effects between sham and experimental conditions.

Indeed, for the group receiving experimental training first, carry-over effects from the first training session to the second were also observed. Specifically, for participants who began with the experimental training, the observed increase in non-harmonic alpha-theta ratios persisted into the subsequent sham training session. This observation suggests that the effects of the experimental training endured, with participants maintaining increased levels of non-harmonic alpha-theta ratios during the sham training similar to those at the conclusion of the experimental training phase. Congruent findings have been observed in other neurofeedback studies, indicating that individuals who attain a specific neural state through neurofeedback training can maintain those performance levels even after the training has ended (Parsons & Faubert, 2021).

### Transferability of Non-Harmonic Alpha-Theta Cross-Frequency Interactions

When assessing transferability, we observed a significant difference in the trajectory of the incidences of the non-harmonic alpha-theta ratio range from the pre-intervention to post-intervention resting-state recordings between the experimental and sham trainings. Similarly as above, the observed effect was only evident at the global (whole-scalp) level but not at the local (trained Pz electrode) level. This effect indicated a significant increase in the incidence of the trained non-harmonic alpha-theta ratio range during the resting-state recording following the experimental training, whereas no significant resting-state changes were observed following the sham training. This finding is in line with the earlier observed whole-scalp training effect throughout the training trials, as it suggests that the training’s impact on the alpha-theta ratio range is a more widespread phenomenon rather than being restricted to the trained electrode location. This finding also implies that the evolution in the incidence of the non-harmonic alpha-theta ratios that was previously observed during the training trials persists into the resting-state recordings and thus outside the active training context. This finding is particularly noteworthy as changes in resting-state brain activity are not always apparent following neurofeedback studies, despite successful training sessions (e.g., Escolano et al., 2014; Navarro Gil et al., 2018; Soriano et al., 2023). Importantly, this observed persistence suggests that the neural cross-frequency state fostered by the experimental neurofeedback training is not only trainable but can also be transferred and maintained, even after the explicit training session has ended.

The observed post-training increase in the incidence of the non-harmonic alpha-theta ratio range also aligns with the previous observation that participants who initially underwent the experimental training continued to display a consistent incidence of the non-harmonic alpha-theta ratio range during the later sham training trials. In essence, the experimental training appeared to induce durable neural changes that persisted not only through the subsequent sham training but also into the post-sham resting-state recording conducted approximately 25 minutes later. Together, this demonstration of longer-lasting neural changes, rather than short-lived ones, following neurofeedback training suggests potential for clinical applications. Indeed, for such applications, it is essential that therapeutic approaches yield long-lasting effects rather than temporary ones, as clinical effectiveness and success depend largely on maintaining positive outcomes over an extended period of time.

### Brain-behavior Associations

Significant brain-behavior associations were identified between the incidence of the non-harmonic alpha-theta ratio range and assessments of depressive mood states and general depressive symptomatology. Specifically, individuals with an increased depressive mood state or higher levels of general depressive symptoms at the start of the training exhibited stronger increases in the incidence of the non-harmonic alpha-theta ratio range post-training. Moreover, these training-induced increases were associated with an improvement in depressive mood. These results have important implications as they suggest that individuals with higher initial depressive symptomatology may benefit more from receiving the current neurofeedback training, as evidenced by both increased neurofeedback trainability and concurrent amelioration in depressive symptoms. These observations align with the findings of Rodriguez-Larios & Alaerts (2021), who linked the occurrence of non-harmonic alpha-theta ratios with lower levels of mind wandering, a construct closely associated with depressive symptomatology (Hoffman et al., 2016). Furthermore, the noted shifts in depressive mood states, coupled with the sustained cross-frequency changes from the training sessions into the resting-state recordings, underscore the potential of neurofeedback-induced neural modifications to yield durable cognitive and behavioral benefits (Rance et al., 2018). Furthermore, the identified relationship between depressive symptoms and post-training improvements suggests that baseline assessments of depressive symptomatology could serve as a valuable indicator for identifying neurofeedback responders and non-responders. Such an indicator could be important for determining inclusion and exclusion criteria for future neurofeedback interventions, ultimately optimizing treatment efficacy and resource allocation (Weber et al., 2020). Finally, no significant associations were found between changes in the non-harmonic alpha-theta ratio range and other self-reported psychological or physiological states. These null findings suggest that the incidence of the non-harmonic alpha-theta ratio range is primarily associated with variations in aspects of depressive symptomatology rather than a broader range of states.

Correlation analyses showed no significant associations between the incidence of the non-harmonic alpha-theta ratio range during the initial resting-state recording and concurrent measures of mindfulness skills (as measured by the MAAS questionnaire). Although not significant, a trend towards a medium-sized negative brain-behavior association with mindfulness skills was noted, indicating a trend towards lower mindfulness skills in individuals with a higher incidence of non-harmonic alpha-theta ratios at baseline (ρ = -.277, *p* = .138, see supplementary Table 1A). Albeit at trend-level, this finding contrasts with previous research showing a heightened occurrence of non-harmonic alpha-theta ratios following mindfulness training, suggesting a positive correlation between these non-harmonic ratios and mindfulness skills (Rodriguez-Larios et al., 2020). Considering that the MAAS questionnaire focuses on the *awareness* aspect of mindfulness, future research should incorporate more comprehensive measures of mindfulness that encompass facets such as non-reactive decentering and non-judgmental acceptance. Such measures would enable a more thorough assessment of whether these additional dimensions correlate more strongly with the reduced cognitive engagement suggested by an increase in non-harmonic alpha-theta ratios.

### Spatial and Ratio Specificity of the Upregulated Alpha-Theta Ratios

Cluster-based permutation testing revealed shifts in alpha-theta cross-frequency ratios following the neurofeedback training. Specifically, there was an observable increase in ratios closely matching the 1.6:1 ratio, typically associated with the cognitive state of focused attention meditation (Rodriguez-Larios et al., 2020; Rodriguez-Larios et al., 2020; Rodriguez-Larios & Alaerts, 2021). This increase was accompanied by a decrease in ratios approximating 2:1, which have been linked to the presence of a cognitive load and the propensity for mind wandering (Rodriguez-Larios & Alaerts, 2019; Rodriguez-Larios et al., 2020). These ratio changes suggest a shift towards a more focused meditative state following the neurofeedback training, indicating a potential quieting of the mind. Our findings further indicated that the effects were most prominent at frontal electrode locations, aligning with previous research that suggests that mindfulness meditation involves neural modulation in prefrontal brain regions, serving as a mechanism for top-down attentional control (Rathone et al., 2022).

### Limitations

The results of our study should be interpreted with the following methodological limitations in mind. Firstly, our experimental training was designed to upregulate a range of cross-frequency ratios approximating the 1.6:1 non-harmonic alpha-theta ratio, rather than targeting this specific ratio itself. Targeting this broader range was adopted to allow for providing an adequate amount of positive auditory feedback throughout the training trials, which is deemed important for successful training. However, future research is needed to examine whether targeting the specific 1.6:1 non-harmonic alpha-theta ratio itself, rather than the adopted range of cross-frequency ratios, may be more efficient for yielding beneficial training effects and transfer. Another limitation of this study is the use of a single-session neurofeedback intervention. Whereas this single-session protocol entailed significant neural changes, the duration of the intervention may have been too brief to observe substantial changes at the Pz electrode alone. Indeed, neurofeedback effects often emerge more clearly over multiple sessions, suggesting that a more extended study design might have revealed more specific training effects (e.g., Domingos et al., 2021). Next, the within-subject design of our study appeared to have introduced carry-over effects from one training to the next, which could have confounded the results. Specifically, participants who first underwent experimental training appear to exhibit a retention of the trained neural state in the subsequent sham session. In contrast, those who experienced the sham training first showed less pronounced responses during later experimental training. These sequence effects suggest that initial sham training experiences may influence the efficacy of remaining neurofeedback trials. Therefore, future research could benefit from a between-subjects design to eliminate such carry-over effects and provide a more accurate assessment of neurofeedback efficacy.

## Conclusion

The current study sought to assess the trainability of non-harmonic alpha-theta cross-frequency ratios, which are known to be associated with focused attention meditative practices, during neurofeedback training. Our findings indicated a successful upregulation of these non-harmonic alpha-theta ratios during a single-session training. Moreover, the observed increases in non-harmonic alpha-theta ratios were linked to reduced depressive mood states, and individuals with higher levels of depressive mood states at baseline exhibited greater trainability. Taken together, these results open new avenues for future research to explore the potential of utilizing extended, multiple-session neurofeedback training programs to enhance meditative practices.

## Author Contributions

Hendrik-Jan De Vuyst: Methodology, Validation, Formal analysis, Investigation, Data Curation, Writing - Original Draft, Writing - Review & Editing, Visualization, Project administration; Angeliki-Ilektra Karaiskou: Methodology, Software, Validation, Formal analysis, Data Curation, Writing - Review & Editing; Javier R. Soriano: Methodology, Software, Formal analysis, Writing - Review & Editing, Visualization; Jelle Pergens: Investigation, Data Curation, Writing - Original Draft, Writing - Review & Editing; Ruth Op de Beeck: Investigation, Writing - Review & Editing; Katleen Van der Gucht: Conceptualization, Methodology, Writing - Review & Editing, Supervision, Funding acquisition; Filip Raes: Conceptualization, Methodology, Writing - Review & Editing, Supervision, Funding acquisition; Maarten De Vos: Conceptualization, Methodology, Writing - Review & Editing, Supervision, Funding acquisition; Carolina Varon: Conceptualization, Methodology, Writing - Review & Editing, Supervision, Funding acquisition; Kaat Alaerts: Conceptualization, Methodology, Validation, Formal analysis, Resources, Data Curation, Writing - Review & Editing, Visualization, Supervision, Project administration, Funding acquisition

## Ethics declarations

### Ethics approval

The study was approved by the ethics committee of UZ/KU Leuven (S68049) and conducted in accordance with the Declaration of Helsinki.

### Consent to participate

Written informed consent was obtained from all individual participants included in the study.

### Data Availability Statement

All data is made publicly available on the Open Science framework (https://osf.io/p93v5/?view_only=b6202233385a4f539b1b43507365a186)

---

**Supplementary Table 1.** Spearman correlation coefficients between baseline emotional distress, mood states, and mindfulness skills and (i) the baseline incidence of the non-harmonic ratio range (A, top panel), as well as (ii) changes in the incidence of the non-harmonic ratio range from pre-to-post-resting state (B, bottom panel).

|  | Baseline Emotional Distress<sup>a</sup> |  |  |  | Baseline Mood State<sup>b</sup> |  |  |  | Baseline Mindfulness Skills<sup>c</sup> |
| --- | --- | --- | --- | --- | --- | --- | --- | --- | --- |
|  | Depression | Anxiety | Stress | Total | Depression | Agitation | Sleepiness | Vigilance | Total |
| A. Baseline incidence of the non-harmonic ratio range | .351 | .112 | .354 | .257 | .041 | .059 | .284 | -.235 | -.277 |
| B. Pre- to post-resting-state changes in the incidence of the non-harmonic ratio range | .376* | .311 | .275 | .345 | .574** | .147 | .297 | -.112 | -.275 |

<sup>a</sup>as measured using the Depression, Anxiety, and Stress Scale (DASS-21)
<sup>b</sup>as measured using the Profile of Mood State (POMS)
<sup>c</sup>as measured using the Mindful Attention Awareness Scale (MAAS)
\*correlation coefficient is significant at the 0.05 level
*\*\*correlation coefficient is significant at the 0.01 level*

**Supplementary Table 2.**
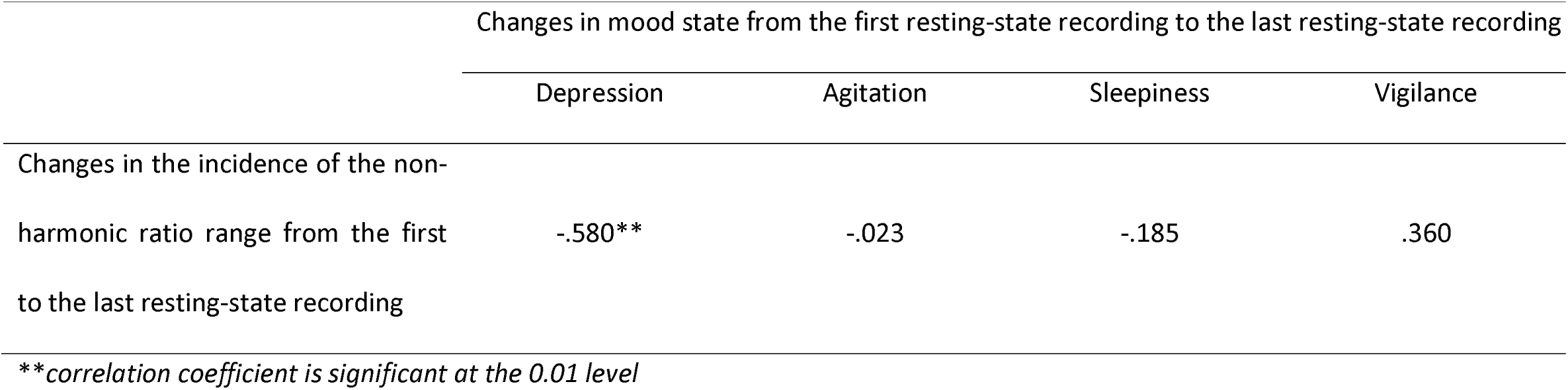
Spearman correlation coefficients between changes in the incidence of the non-harmonic ratio range from the first resting-state recording to the last resting-state recording, and changes in concurrently assessed mood states.

|  | Changes in mood state from the first resting-state recording to the last resting-state recording |  |  |  |
| --- | --- | --- | --- | --- |
|  | Depression | Agitation | Sleepiness | Vigilance |
| Changes in the incidence of the non-harmonic ratio range from the first to the last resting-state recording | -.580** | -.023 | -.185 | .360 |

— \*\*correlation coefficient is significant at the 0.01 level

**Supplementary Table 3.**
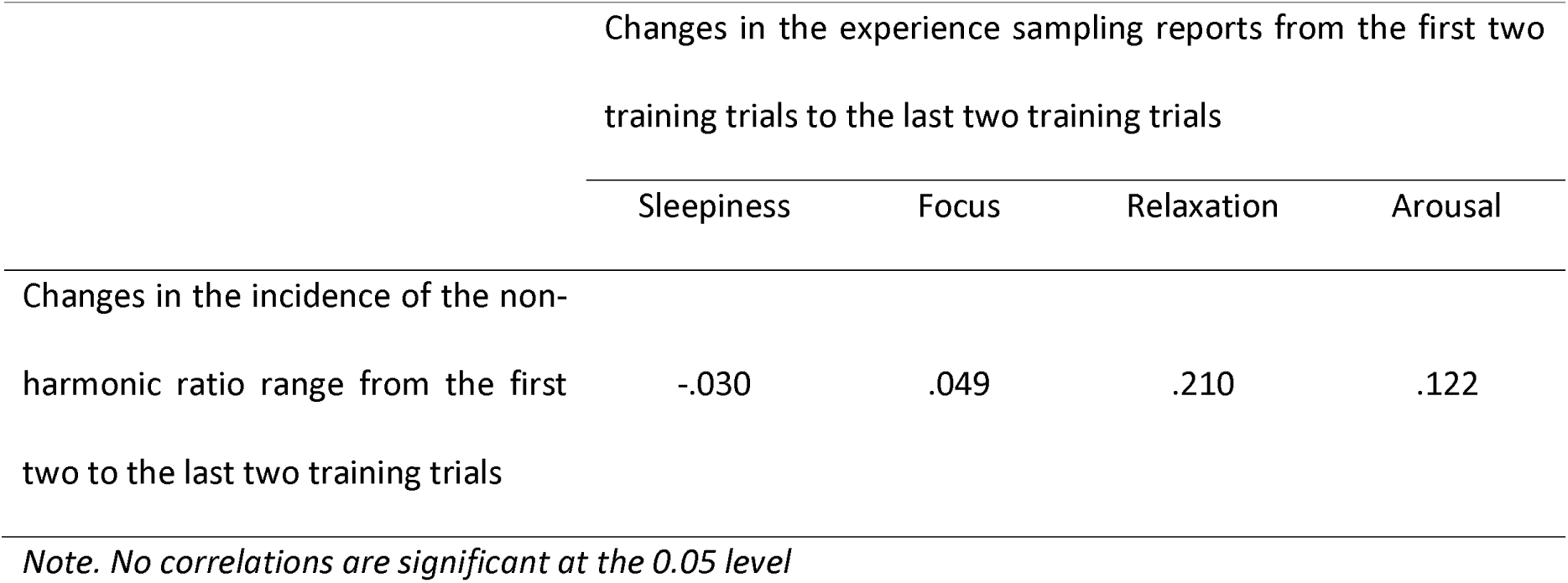
Spearman correlations coefficients between changes in the incidence of the non-harmonic ratio range from the first two training trials to the last two training trials, and changes in the concurrent experience sampling reports.

